# Impact of an AI medical scribe after 375 000 notes generated across care levels in a European health system

**DOI:** 10.64898/2025.12.06.25341757

**Authors:** Enni Sanmark, Ville Vartiainen, Johan Sanmark, Katarina Wettin, Lukas Saari, Artin Entezarjou

## Abstract

**Importance:** Reducing clinicians’ documentation burden is a critical priority in modern health care, as excessive administrative work consumes substantial clinician time, contributes to burnout, and limits time available for direct patient care.

**Objective:** To quantify the impact of an AI medical scribe on documentation time and clinician experience.

**Background:** Clinicians spend a substantial share of their working hours on documentation, contributing to workflow inefficiencies, reduced patient-facing time, and increased burnout. AI medical scribes have emerged as a promising solution to reduce this burden, yet real-world evidence remains limited and heterogeneous. Data from European health systems are especially scarce, despite growing interest in AI-enabled documentation support.

**Exposure:** Use of AI medical scribe.

**Design, Setting and Participants:** This observational real-world evaluation was conducted between April 26th 2024 and October 27th 2025 to assess the impact of an AI medical scribe on documentation time and clinician experience using retrospective paired ratings. The study was carried out across multiple specialties in primary, secondary and hospital care within Capio Ramsay Santé, a large integrated health care provider operating in Sweden.

The target population consisted of licensed clinicians actively using the AI medical scribe in routine clinical practice. Eligibility was limited to “fully onboarded” users, defined as clinicians who had used the scribe for at least 3 months, created more than 100 notes, generated at least one document or certificate, and used the conversational edit (“Add or adjust”) feature at least once.

**Results:** With the introduction of the AI medical scribe, the estimated time spent on documentation per note decreased from 6.69 minutes to 4.72 minutes (-29%, p = 1.70e-11). On a five-point Likert scale, the ability to work without stress related to administrative tasks increased from a mean of 2.41 to 3.14 (p = 2.46e-8), and perceived presence with patients increased from 3.73 to 4.33 (p = 2.47e-8). The median editing time was 93 seconds, and it did not decrease significantly over continued use.

**Conclusions and Relevance:** This study shows that the clinician time savings and reductions in cognitive load and stress reported in prior US-based studies can also be achieved in a European health care system using an AI scribe.

**Key Points:** **Question:** Does implementation of an AI medical scribe reduce subjective documentation time and improve clinician experience in European health care settings?

**Findings:** In this observational real-world evaluation of licensed health care professionals using an AI medical scribe within a large Swedish health care provider, the estimated time spent on documentation decreased from 6.69 to 4.72 minutes (-29%) per note, and clinicians reported statistically significant improvements both in their ability to work without administrative-task–related stress, and in their perceived presence with patients. Median note-editing time was 93 seconds and did not change significantly with continued use.

**Meaning:** These findings suggest that AI medical scribes can meaningfully reduce documentation burden and cognitive load for clinicians, potentially freeing more time and attention for direct patient care.

## 1 Introduction

Integrating artificial intelligence (AI) into health care has created substantial opportunities to improve clinical efficiency but also enhance patient care, and increase clinicians’ professional satisfaction. One of the most compelling possibilities lies in reducing clinicians’ documentation burden. Although electronic health records (EHRs) have contributed to improved health outcomes ^1^ and reduced medical errors ^2^ they have also become a dominant part of clinicians’ workdays and are linked to diminished job satisfaction and increased burnout ^3, 4^ ^5^Research consistently shows that clinicians spend up to half of their working hours on computers, primarily performing documentation tasks ^6^^7 8 9^, while less than 30% of time is devoted to direct patient care^10^. In this context, ambient large-language-model-based AI medical scribes able to transcribe clinician–patient interactions and generate structured draft notes have emerged as a promising and scalable means to substantially reduce documentation load. Thereby, they may be able to free time, improve workflow, support better patient care, improve quality of notes and ultimately strengthen clinician well-being across specialties and care settings ^11 12 13 14 15^

Evidence on the effectiveness of AI-assisted clinical documentation remains limited, heterogeneous, and in part contradictory. Early large-scale deployments, primarily in North America, suggest that AI medical scribes can reduce time spent on documentation, decrease after-hours electronic health record (EHR) work, and improve elements of clinician–patient interaction ^12,13 14 16^ . However, reported benefits vary markedly across clinical specialties, workflows, and implementation strategies, emphasizing the need for context-specific evaluation ^12 14^. Conversely, some studies have found no measurable improvements in efficiency, either in terms of time savings or economic outcomes ^17^. Complicating matters further, the underlying technology is evolving at a rapid pace making it challenging to draw stable conclusions about current real-world performance. Despite these uncertainties, health care organizations are making adoption decisions now, and AI scribe systems are already in use in nearly all major U.S. health systems ^18^. This rapidly evolving landscape underscores the importance of rigorous, context-sensitive evaluation of their effectiveness.

European health systems operate within regulatory and operational conditions that may substantially influence the performance and adoption of AI medical scribe technology (MDCG 2019-11; EU MDR 2017/745). Combined with Europe’s heterogeneous EHR landscape and multilingual clinical environment, these factors create a context that differs markedly from non-EU settings ^19^. Evidence from European countries remains especially limited ^2021^, despite their high digital maturity and increasing interest in reducing administrative burden and improving clinician experience through advanced automation.

The aim of this study is to evaluate the impact of a fully deployed AI medical scribe on clinical documentation time and clinician experience in primary care and secondary care (including hospital care) across multiple specialties. Together, these outcomes provide a comprehensive assessment of the effectiveness and usability of AI-scribe technology in Europe during large-scale implementation in routine clinical practice.

## 2 Methods

### 2.1 Participants, Setting and Exposure

This observational study was conducted between April 26th 2024 and October 27th 2025 in health care facilities run by Capio - part of Ramsay Santé (Capio Headquarters, Gothenburg, Sweden). The study uses two data sources. Firstly, metadata was gathered from all participating units which implemented the same CE-marked AI medical scribe (Tandem AI Medical Scribe, version 1.0 - 1.1, manufactured by Tandem Health AB., Stockholm, Sweden). Secondly, a survey was conducted among fully onboarded professionals to gather subjective assessments on the use of the scribe.

The study does not constitute human participants research, as it involved analysis of deidentified, aggregated survey data for which ethical approval is not required. Informed consent was obtained from all survey respondents. Survey responses were linked to exported metadata on secure infrastructure under organizational data-governance policies. Metadata export from the vendor’s system included only the elements required for the preregistered analyses (timestamps, counts, and contextual descriptors).

The author responsible for the statistical analysis (V.V.) was not involved in the intervention, had no contact with participants, and received no incentives or remuneration from the vendor. The study adhered to the Standards for Quality Improvement Reporting Excellence (SQUIRE) guideline and was preregistered on the Open Science Framework on 7 October 2025 (DOI: 10.17605/<u>OSF.IO/YPD9E</u>)

Primary outcome was defined as change in self-reported time used to complete a typical clinical note with and without the scribe. Secondary outcomes were change in responses to questions “I can work without feeling stressed by administrative tasks” and “I feel fully present with the patient during the consultation” in five-point Likert scale. Additional outcomes were yes/no responses to questions related to user satisfaction related statements.

The exposure consisted of routine use of the AI medical scribe within the health care provider’s clinical operations following standard implementation. Deployment included a structured onboarding program delivered by clinician-informed customer success staff from the vendor: an initial 2-hour foundational session covering system introduction, first note generation, and transfer of the note to the EHR, followed by two 1-hour reinforcement sessions at approximately 4 and 8 weeks to review advanced features and address local workflow needs.

Additionally, the health care provider held a 1-hour session with management and senior clinicians focusing on responsible change management, as well as a separate 1-hour course for all end-users focusing on how to use the system responsibly and safely with patients in clinical practice. Adoption at each site was then supported by designated superusers, while the centralized IT organization ensured installation, integration, and basic operability across all participating clinics.

#### 2.1.1 Survey

The survey was sent to all fully onboarded clinicians by email with two additional reminders when necessary. The inclusion criteria were: (i) used the scribe for ≥3 months, (ii) created >100 notes with the scribe, (iii) generated at least one document/certificate, and (iv) used the conversational edit (“Add or adjust”) feature at least once. The complete survey is presented in supplementary material S1.

Time was assessed as the self-reported estimated number of minutes required to complete a typical clinical note with and without the scribe. The survey also included paired 5-point Likert scale items to measure perceived administrative burden (“I can work without feeling stressed by administrative tasks”) and clinician presence (“I feel fully present with the patient during the consultation”) with and without the scribe. Additional survey items were yes/no responses to questions related to user satisfaction related statements “Tandem is easy to use”, “I want to continue using Tandem”, and "Would you recommend Tandem to colleagues in your or other organizations?”

#### 2.1.2 Note metadata

Each note contained pseudonymized linking to the professional for whom the note was generated as well as the clinical site. Editing time per note was estimated from the automatically generated metadata and was defined as the number of seconds between the timestamp of the first edit and the timestamp of the last edit for each completed note.

Collected variables and definitions are specified below:

● **Profession:** Each note is associated with a unique clinician user account. Clinicians access the AI medical scribe using strong electronic identification (a health care professional smart card or a government-issued electronic ID application linked to their national personal identity number), which ensures that each account corresponds to a specific licensed individual. For each note, the clinician’s profession was taken from the account-level role metadata and treated as the note-level profession variable. Notes generated by users with a tagged profession of “clinic admin” or “medical secretary” were considered non-clinical activity and were excluded from the analytic cohort.
● **Visit type:** The visit type was derived from the template used when generating the note. Clinicians actively choose a note template at the start of documentation, and templates are designed and maintained by the health care provider and vendor’s operations team, who know for which clinical context each template is intended. Each distinct template name was thus mapped to one of four encounter categories: physical (in-person visits and procedures), digital (remote written/online contacts), telephone, or admin/other, with unmatched templates falling into the admin/other category.
● **Platform:** Clinicians could log in either via a native desktop application or via a web browser on their desktop computer, or on their mobile devices. The platform could thus capture what access method was associated with each note, enabling categorization to desktop or mobile access.

### 2.4 Statistical analysis

All statistical analyses were performed in R (version 4.3.3, R foundation for statistical computing, Vienna, Austria).

Two complementary datasets were analyzed: (1) note-level editing metadata and (2) clinician-level survey responses describing use of the AI-assisted documentation system.

Editing time was defined as the time elapsed from when the professional pasted the text to when they made the final modification. Editing durations displayed a pronounced right-skew with extreme values attributable to system artifacts (e.g., notes left open for prolonged periods). To mitigate undue influence of these outliers, we applied a 3-hour cutoff, excluding notes with edit durations >10,800 seconds. Edit times were further log-transformed to stabilize variance and reduce skewness. To examine within-user changes in editing time, we compared each user’s first 2 months of recorded note activity with the remainder of the study period.

To evaluate factors associated with editing duration at the note level, we fit a linear mixed-effects model with log-transformed edit time as the dependent variable. Fixed effects included visit type, profession, clinic type, and interaction platform.

A random intercept for clinician was included to account for within-clinician correlation arising from repeated notes. Model assumptions were assessed visually via Q–Q plots of residuals and residual-versus-fitted plots. This graphical evaluation indicated modest deviations from normality in the tails, which is expected with large-scale operational data, but did not materially affect inference.

Since Likert-type responses and recall measures were not normally distributed, paired Wilcoxon signed-rank tests were used to compare editing time recall (before vs. after), perceived stress (without vs. with AI medical scribe), and perceived presence (without vs. with AI assistance). To examine whether changes in editing behavior were associated with subjective experience, we fit linear regression models with Δ stress, Δ presence, or Δ recall as the outcome and Δ log-edit time as the primary predictor. For completeness, sensitivity analyses using median edit times instead of mean log-edit times produced consistent conclusions. For yes/no survey items, the proportion answering ‘yes’ was calculated, and 95% confidence intervals were obtained using the exact binomial method.

## 3 Results

The original data set contained metadata from 376,596 clinical notes. The most active user had generated 2,474 clinical notes. After the filtering and removal of artefacts the final dataset contained 236,153 unique notes across 1,295 participants. We excluded 138,196 notes with no recorded edits and 2,247 notes with editing times exceeding three hours. The cumulative number of notes and implementation periods are presented in Figure 1.

**Figure 1:**
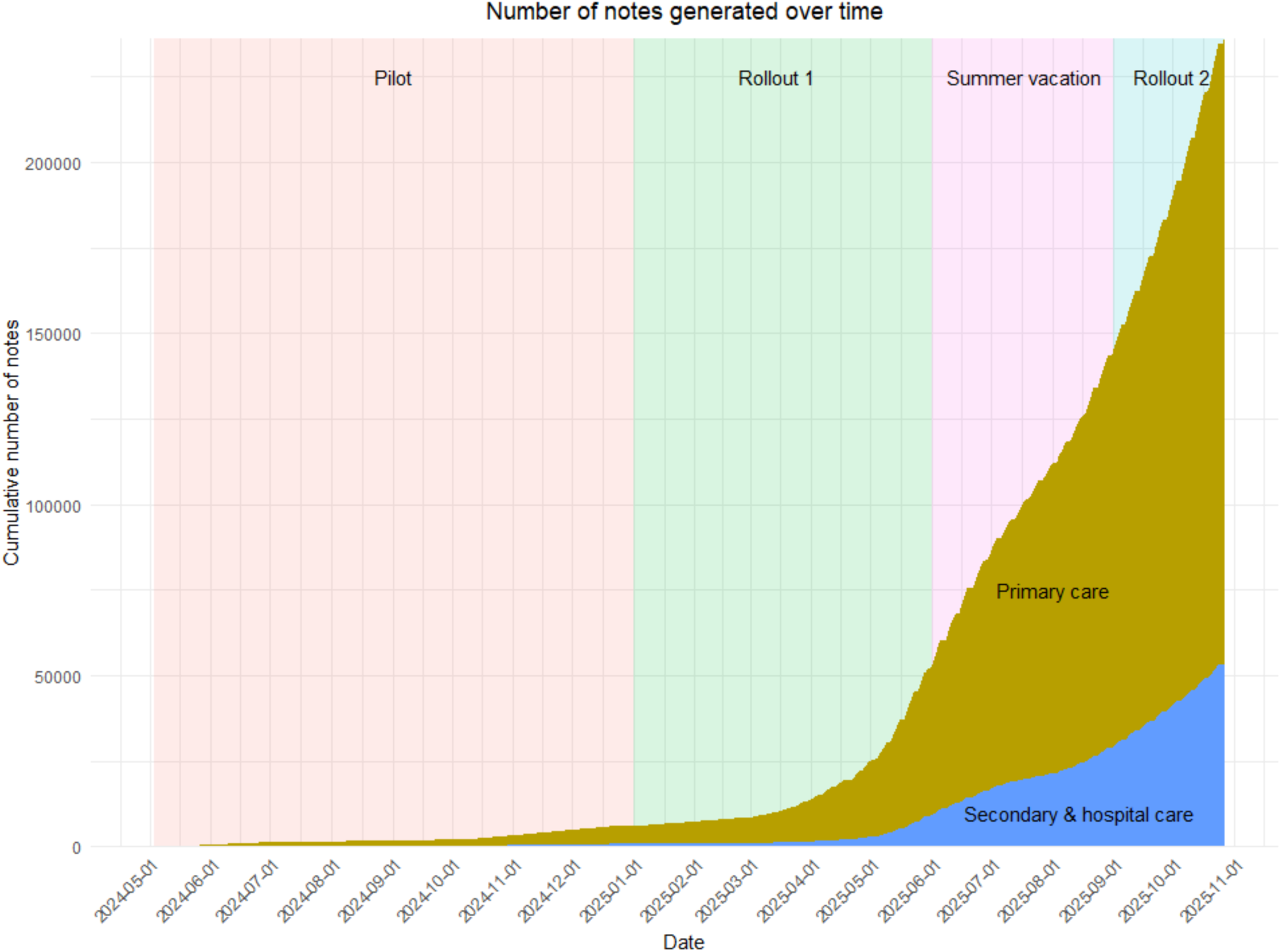
Cumulative number of notes generated during different stages of implementation in primary care and secondary care (including hospital care).

The three most common professions were general practitioners (65.9%), physiotherapists (6.1%) and orthopedic surgeons (5.1%). The remaining 22.9% of participants represented a wide variety of professions including various medical specialities, nurses, dentists, and optometrists (Fig. S1). A total of 177 participants, who together created 67,679 notes, responded to the survey.

### 3.1 Metadata-related results

The median editing time was 93 seconds, whereas mean editing time was 226 seconds after applying the designated artifact cut-off of 3 hours, indicating a highly skewed distribution. The median remained stable in the sensitivity analysis: in the full dataset of generated notes, the median was 94 seconds, and with a 30-minute cut-off, the median was 87 seconds. When notes were stratified into those generated within the first two months of use versus those generated thereafter, the median editing times were 84.0 s and 84.5 s, respectively, suggesting that no substantial learning curve was observed (Fig. 2)

**Figure 2.**
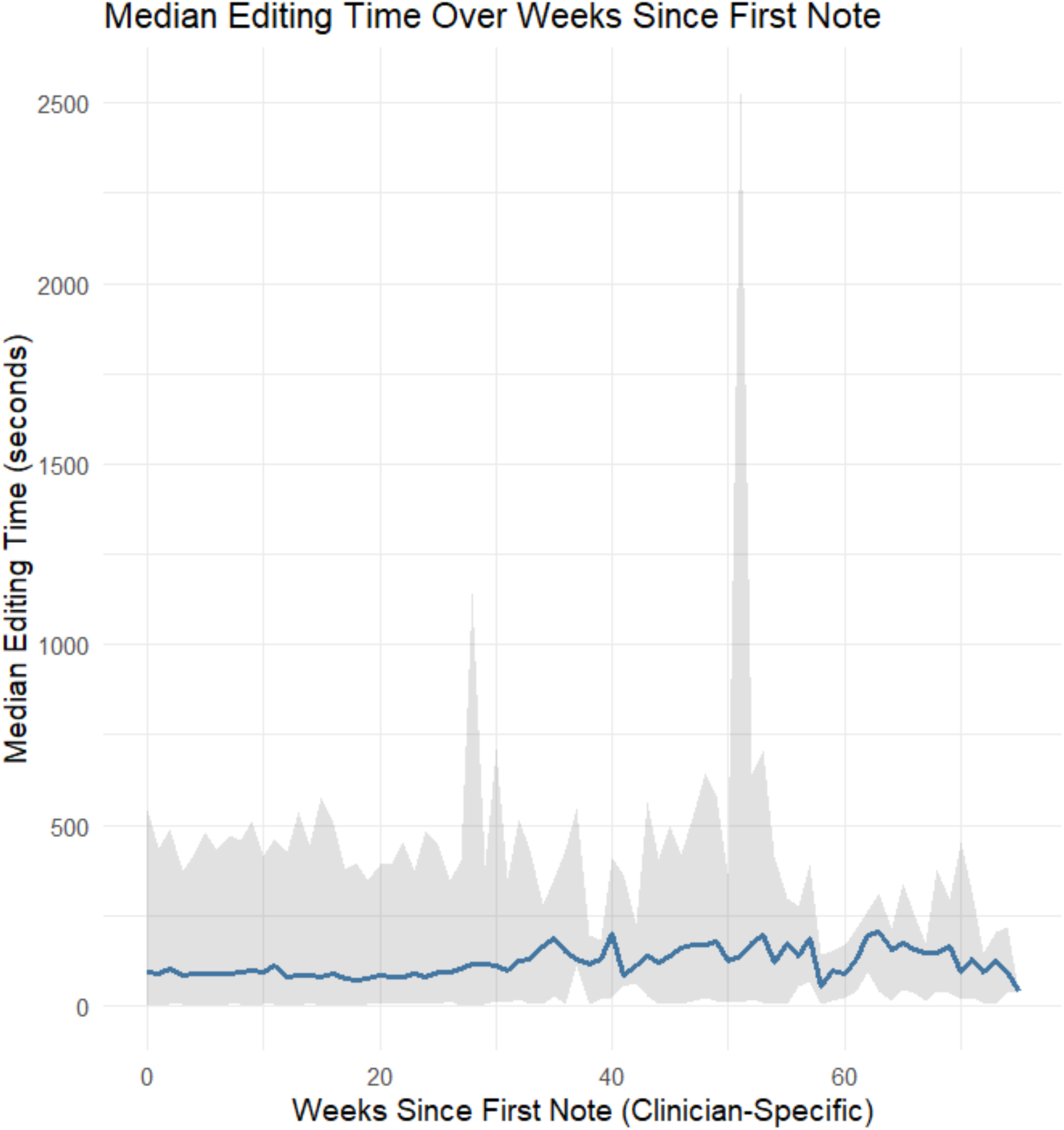
Median editing time as a function of time. The time is defined as weeks from the onboarding for each user separately. The curve shows the weekly medians across clinicians, with the shaded area indicating variability (2.5th–97.5th percentiles).

The effect of visit type and platform on editing time were studied using a generalized linear mixed-effects model, as presented in table 1. Differences between professions were minimal and statistically non-significant and were therefore excluded from the final model.

**Table 1.** Linear mixed model for log transformed edit time.

| Variable | $\beta$<br>[($e^\beta-1$) · 100 %] | p-value | 95% CI<br>[($e^\beta-1$) · 100 %] |
| --- | --- | --- | --- |
| **Visit type** |  |  |  |
| **Physical** | Reference |  |  |
| **Digital** | -0.9599<br>[-61.7%] | <2e-16 | -1.080 — -0.840<br>[-66% -57%] |
| **Telephone** | -0.7988<br>[-55.0%] | <2e-16 | -0.872 — -0.726<br>[-58% -52%] |
| **Admin/other** | -1.060<br>[-65.3%] | <2e-16 | -1.097 — -1.024<br>[-67% -64%] |
| **Platform** |  |  |  |
| **Desktop** | Reference |  |  |
| **Mobile** | 0.7010<br>[+102%] | <2e-16 | 0.580 — 0.822<br>[] |

*CI = confidence interval, [( $e^\beta-1$ ) · 100 %] gives relative time difference to reference*

**Table 2.**
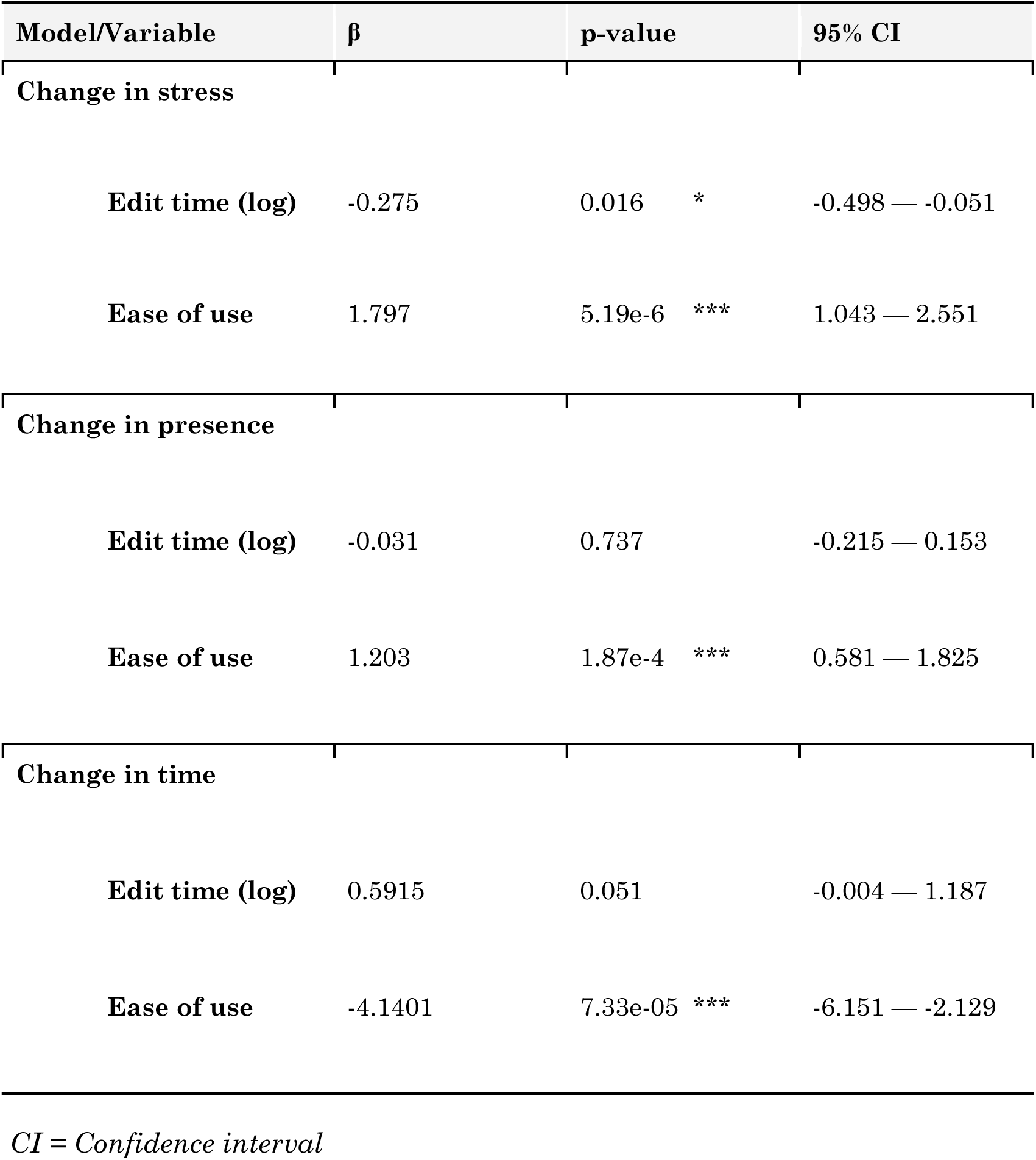
Linear regression models for changes in ability to work without stress of administrative tasks, presence with patients and perceived time used for documentation.

| Model/Variable | $\beta$ | p-value | 95% CI |
| --- | --- | --- | --- |
| **Change in stress** |  |  |  |
| **Edit time (log)** | -0.275 | 0.016 * | -0.498 — -0.051 |
| **Ease of use** | 1.797 | 5.19e-6 *** | 1.043 — 2.551 |
| **Change in presence** |  |  |  |
| **Edit time (log)** | -0.031 | 0.737 | -0.215 — 0.153 |
| **Ease of use** | 1.203 | 1.87e-4 *** | 0.581 — 1.825 |
| **Change in time** |  |  |  |
| **Edit time (log)** | 0.5915 | 0.051 | -0.004 — 1.187 |
| **Ease of use** | -4.1401 | 7.33e-05 *** | -6.151 — -2.129 |

*CI = Confidence interval*

### 3.2 Survey-related results

In the predefined primary outcome, the estimated time spent on documentation decreased from 6.69 minutes to 4.72 minutes (Fig. 3, p = 1.70e-11), and on a five-point Likert scale (Fig. 4), the ability to work without stress related to administrative tasks increased from 2.41 to 3.14 (p = 2.46e-8,) and perceived presence with patients increased from 3.73 to 4.33 (p = 2.47e-8).

**Figure 3.**
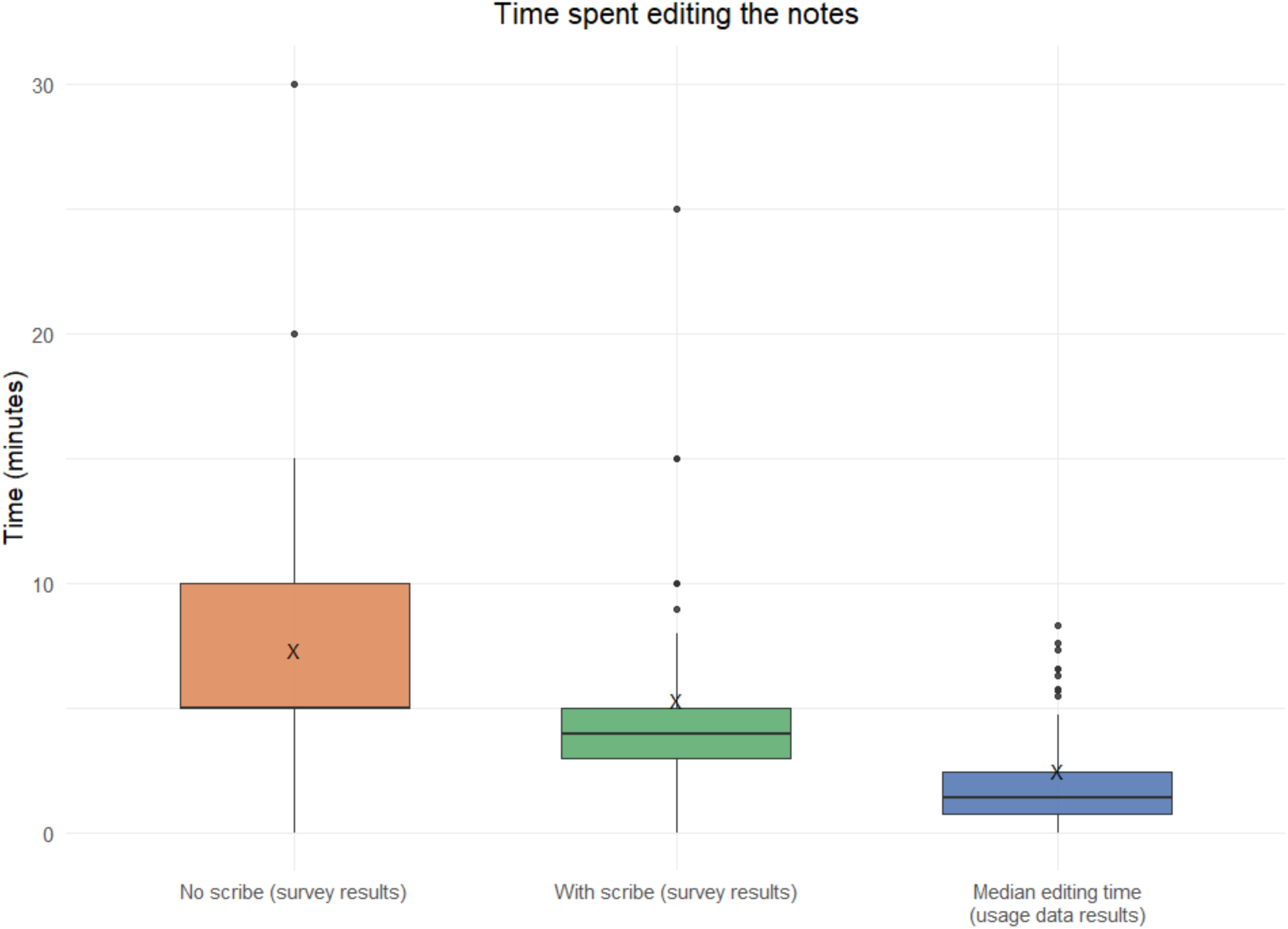
Self-estimated time to complete a single clinical note with and without the scribe as well as objectively measured median editing time for each clinician. X denotes the mean.

**Figure 4.**
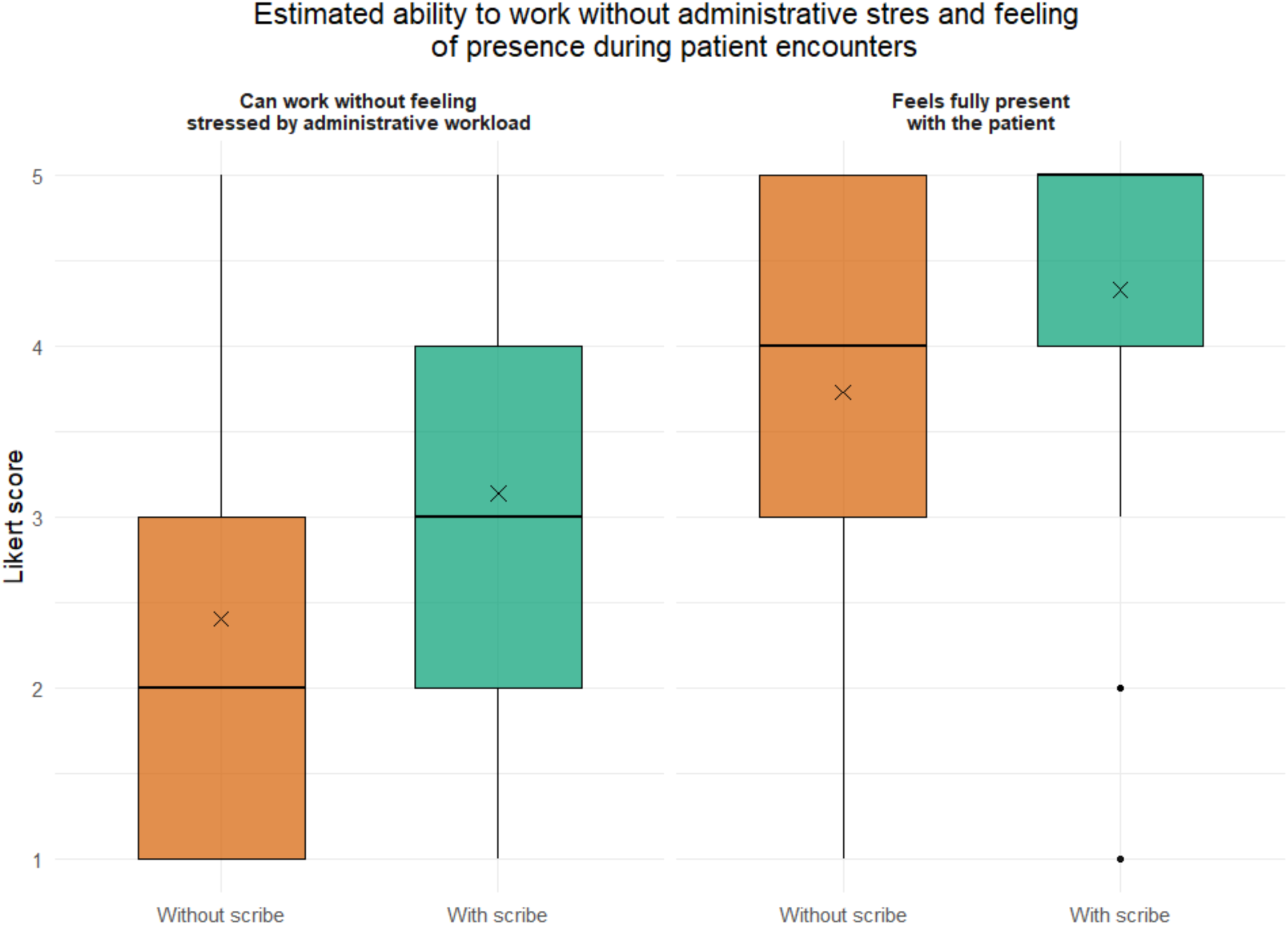
Likert scale answers to questions assessing ability to work without stress from administrative tasks and feel fully present with the patient with and without AI scribe. X denotes the mean of the answers.

In the survey items indicating yes/no answer 80.3% (95% CI 73.6-85.99%) said the scribe was an improvement for their profession; 91.3% (86.1-95.1%) found it easy to use; 91.3% (86.1-95.1%) wished to continue using the scribe; and 84.4% (78.1-89.6%) would recommend colleagues at other clinics to start using the scribe.

Three linear regression models examined whether objectively measured note-editing time (log-transformed) and perceived ease of use predicted self-reported changes in documentation time, administrative stress, and patient presence. Lower objective editing time was significantly associated with reduced administrative stress (ꞵ = -0.275, p = 0.016, 95% CI -0.498, -0.051), but it did not significantly predict perceived documentation time (ꞵ = 0.592, p = 0.051, 95% CI -0.004, 1.187), or presence with patients (ꞵ = -0.031, p = 0.737, 95% CI -0.215, 0.153).

In contrast, perceived ease of use was a significant predictor in all models: higher ease-of-use ratings were associated with reduced administrative stress (ꞵ = 1.797, p < 0.001, 95% CI 1.043, 2.551), increased patient presence (ꞵ = 1.203, p < 0.001, 95% CI 0.581, 1.825), and lower perceived documentation time (ꞵ = -4.140, p < 0.001, 95% CI -6.151, -2.129).

## 4 Discussion

### 4.1 Principal findings

Previous studies have demonstrated that AI medical scribes can reduce perceived documentation time as well as stress, thereby potentially lowering the risk of burnout among clinicians ^15^. However, these findings have almost exclusively originated from the United States, whose health care system differs substantially from European systems in terms of organization, financing, and clinical workflows ^12,13 14 15^. In our study, we showed that, in a European context, the use of an AI scribe was similarly associated with reduced physician-reported stress and a decrease in self-reported time spent on documentation.

Our findings on perceived time savings are consistent with the majority of published studies. However, not all studies have been able to demonstrate a reduction in documentation time ^17^. In most prior work, time has been assessed subjectively, as in our study, but objective time measurements reported in some studies also support the observation that medical scribes can reduce the time burden of documentation ^14^.

Previous studies have additionally reported differences between professional groups in the time spent in clinical documentation^7^ . In contrast, we did not detect meaningful differences between professions in our analysis, suggesting that the potential advantages of AI medical scribe use may generalize across clinician groups in this setting.

Secondly, Liu et al. observed that the extent of clinicians’ use of an AI medical scribe influenced the magnitude of benefit they derived in their daily work^17^. In contrast, in our study we did not observe any substantive change in editing time between the initial phase of use and subsequent periods, suggesting that efficiency gains—at least with respect to note editing time—are attained early, remain comparatively stable over time, and are indicative of a technology that is readily usable without a substantial learning period.

In our study, the time required to edit notes generated by the AI medical scribe was relatively short, with a median of 93 seconds. This may partly reflect the structured onboarding provided to users (approximately five hours), which could have increased familiarity with the interface and editing workflow. Notably, a subset of editing episodes extended over several hours. Because editing time was defined as the interval between the first and last recorded edit, periods of inactivity were not separable from active editing.

Prolonged intervals may therefore reflect a common workflow in which the editing window is left open and the note is finalized later, rather than sustained continuous editing.

Both visit type (in-person, digital, telephone) and editing platform (desktop vs mobile) were associated with variation in editing time. Remote encounters resulted in approximately 62% and 55% less editing time for digital and telephone contacts, respectively. On mobile platforms, editing times were roughly double those on the desktop platform. Editing on a desktop interface was also linked to greater editing efficiency. Given that improved documentation efficiency is one of the main drivers for adopting AI medical scribes, these findings highlight the importance of careful selection and optimisation of the user interface. Without attention to workflow design and platform choice, the full potential benefits of the technology may not be realised in routine clinical practice.

Interestingly, physicians’ self-reported documentation time was substantially longer than the measured note editing time. This suggests that, within the time they attribute to “documentation,” clinicians are also performing other tasks, such as writing prescriptions or referrals requiring navigation in the EHR. The estimated time is, however, broadly in line with a previous study ^12^. Conversely, prior work indicates that although health care professionals systematically overestimate the time spent on documentation, often by as much as twofold ^22^, there remains a moderate correlation between actual and estimated time, supporting the use of clinician self-reports as a pragmatic surrogate measure in large-scale process improvement and research ^23 24^. However, this does not remove the need to evaluate time savings and the impact of AI-based medical scribes using a broader set of metrics that capture the full scope of clinical documentation work. The significant number of notes excluded from the analysis due to no editing likely had varying fates such as used as is without editing, transferred and edited in the medical record system or were disregarded.

One important dimension of effectiveness is health care professionals’ experience, which is particularly critical in the context of worsening shortages and increasing burnouts of medical personnel ^25^ ^26^The urgency of addressing clinician well-being is also reflected at the policy level: for example, the National Academy of Medicine convened a meeting in December 2024 to examine the potential for AI to improve health worker well-being, including reducing burnout ^27^. This growing international interest highlights how AI-based tools, such as AI medical scribes, are increasingly viewed not only as instruments for improving efficiency, but also as potential levers to mitigate the psychological burden of clinical work.

In our study, the reduction in stress was associated both with the perceived ease of use of the AI scribe and with shorter editing time for the generated notes. Although these findings could partly be explained by simpler clinical cases requiring less editing, the existing literature do not support the point of view ^13,28^ and it is more likely that they reflect a genuine benefit from automation, leading to a reduced self-assessed workload. This suggests that, beyond potential gains in efficiency, AI medical scribes may contribute meaningfully to improving clinicians’ day-to-day work experience, which is a key outcome in its own right. This interpretation is in line with previous studies reporting that AI medical scribes can reduce clinicians’ cognitive load and, consequently, alleviate stress and the risk of burnout ^15^.

### 4.2 Limitations

The main strength of this study is the large number of medical notes analysed, which likely provides a reliable representation of current editing times in routine clinical practice.

However, we were unable to determine whether the edits made by professionals were clinically necessary for medical quality or patient safety, or whether they primarily reflected stylistic preferences, tone adjustments or other experiential aspects of documentation. The most important limitations relate to the study design. Although this was a mixed-methods study, the survey component was not controlled: it was administered only once and did not include a comparison group. In addition, the survey sample consisted of users who had accumulated sufficient use of the AI medical scribe, which likely over-represents professionals who were more satisfied or at least willing to continue using the tool, introducing potential selection bias.

As in almost all prior work in this area, time savings could not be quantified fully objectively. The editing time has several potential sources of error. While current software allows relatively precise estimation of time spent using the AI medical scribe (e.g. editing time), no baseline time measurements of documentation without the tool were available. Editing time therefore demonstrates that AI medical scribe technology is already capable of supporting efficient documentation, but it does not on its own provide an absolute estimate of time saved. To more robustly assess effectiveness, future studies should incorporate objectively measured baseline and follow-up documentation times, include the full spectrum of users (including those who discontinue or are dissatisfied), and evaluate patient experience, ideally within a randomised controlled study design.

## 5 Conclusion

In our study, we demonstrated that the time savings perceived by clinicians, as well as the reductions in cognitive load and stress reported in previous US-based studies, can also be achieved within a European health care system when using an AI medical scribe.

## 8 Author contributions

● ES: Writing – original draft; Writing – review & editing.
● VV: Formal analysis; Methodology; Validation; Visualization; Writing – original draft; Writing – review & editing.
● JS: Project administration; Writing – review & editing
● KW: Conceptualization; Investigation; Resources; Validation; Writing – review & editing.
● LS: Conceptualization; Methodology; Writing – review & editing
● AE: Conceptualization; Data curation; Formal analysis; Methodology; Project administration; Software; Supervision; Validation; Visualization; Writing – original draft; Writing – review & editing.

## Data Availability

All data produced in the present work are contained in the manuscript

## 9 Supplementary material

S1 Full survey instrument (translated from Swedish). S2 Supplementary figures & tables

### Supplementary material

S1 Full survey instrument (translated from Swedish)

Short survey for the evaluation of the AI assistant Tandem for medical record drafts

We would like to know what differences you experience after having started with Tandem, compared to your situation before gaining access to Tandem. The survey consists only of the questions below and takes approx. 2 minutes to answer.

1. Have you attended at least one onboarding meeting with Tandem’s staff where they explain how the solution works? *Required to answer. Single line text.*

- Yes
- No
2. Which method did you use for clinical documentation before you used Tandem? Required to answer. *Required to answer. Single line text.*

- Dictation to a secretary (transcriptionist)
- Speech recognition (SR, speech-to-text)
- Keyboard (typing)
- ther AI documentation tool
- ther methods for clinical documentation
3. Before you gained access to Tandem: How many minutes did you normally spend finalizing a typical clinical note? *Required to answer. Single line text. The value must be a number*
4. After you have started using Tandem: How many minutes do you normally spend finalizing a typical clinical note? *Required to answer. Single line text. The value must be a number*
5. I can work without feeling stressed by administrative tasks. *Required to answer.*

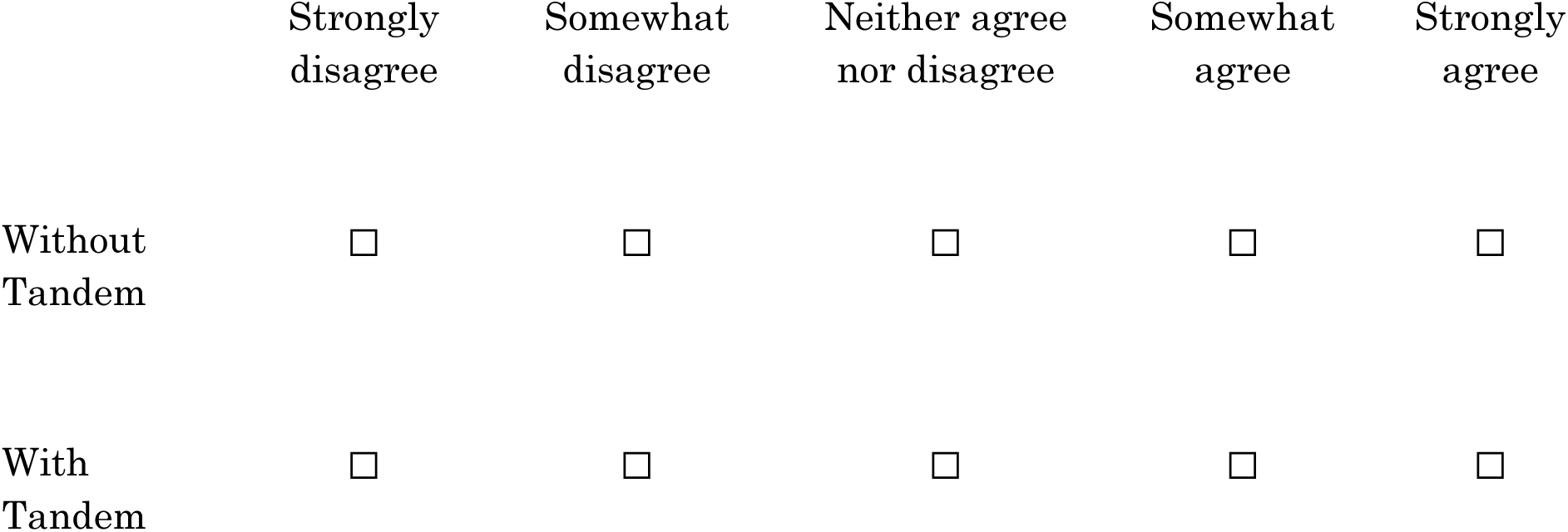
6. feel fully present with the patient during the consultation. *Required to answer.*

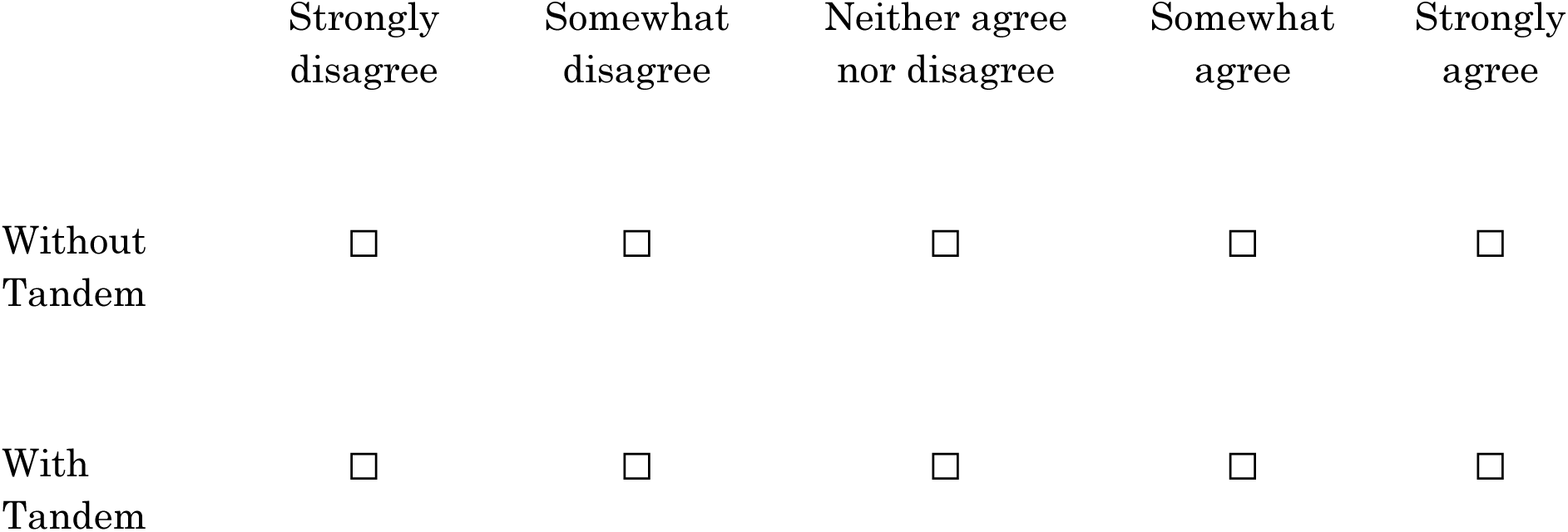
7. Compared to your previous way of working – does Tandem represent an improvement for your profession? *Required to answer. Single choice.*

- Yes
- No
8. Tandem is simple to use. *Required to answer. Single choice.*

- Yes
- No
9. I want to continue using Tandem. *Required to answer. Single choice.*

- Yes
- No
10. Do you recommend colleagues in your unit or at other units to use Tandem? *Required to answer. Single choice.*

- Yes
- No

S2 Supplementary figures & tables

**Figure S1.**
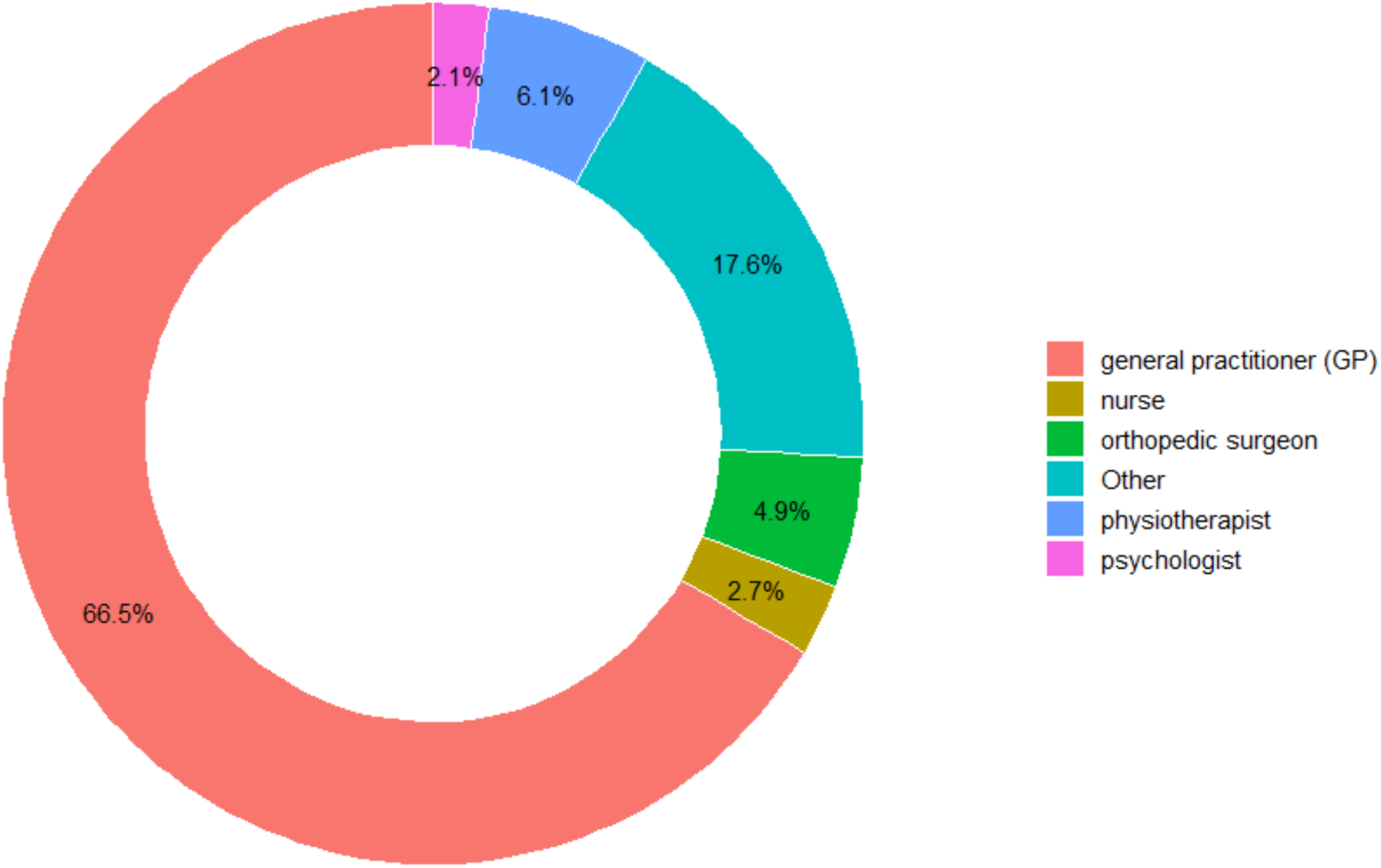
Proportion of professions included in the study. The professions in “other” consist of less than 2% of the total professionals per profession.

**Figure S2.**
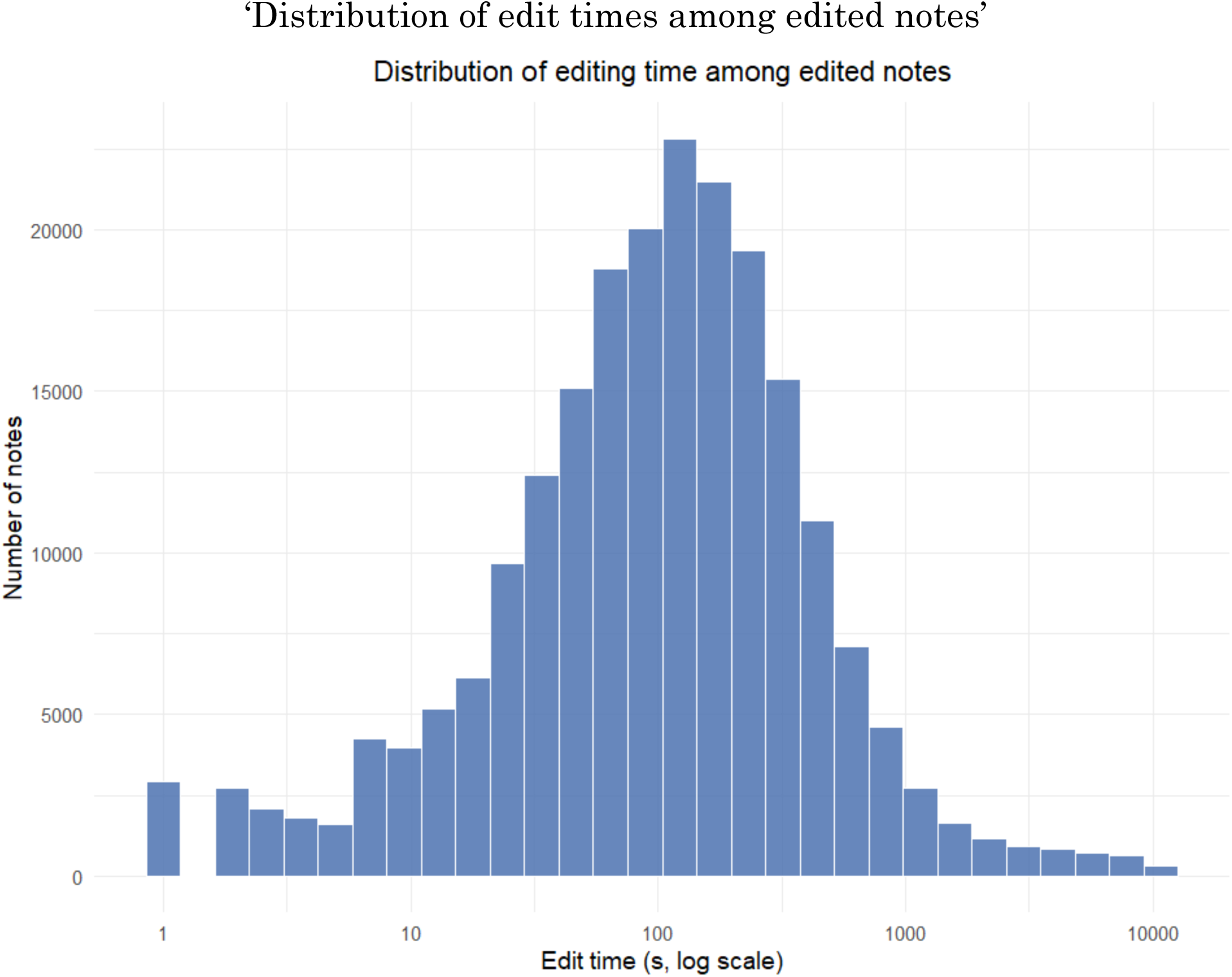
Histogram with log-scaled axis.

