## Supplementary material for "Impact of an AI medical scribe after 375 000 notes generated across care levels in a European health system"

### S1 Full survey instrument (translated from Swedish)

#### *Short survey for the evaluation of the AI assistant Tandem for medical record drafts*

We would like to know what differences you experience after having started with Tandem, compared to your situation before gaining access to Tandem. The survey consists only of the questions below and takes approx. 2 minutes to answer.

1. Have you attended at least one onboarding meeting with Tandem's staff where they explain how the solution works? *Required to answer. Single line text.*

- Yes
- No

2. Which method did you use for clinical documentation before you used Tandem? Required to answer. *Required to answer. Single line text.*

- Dictation to a secretary (transcriptionist)
- Speech recognition (SR, speech-to-text)
- Keyboard (typing)
- Other AI documentation tool
- Other methods for clinical documentation

3. Before you gained access to Tandem: How many minutes did you normally spend finalizing a typical clinical note? *Required to answer. Single line text. The value must be a number*

4. After you have started using Tandem: How many minutes do you normally spend finalizing a typical clinical note? *Required to answer. Single line text. The value must be a number*

5. I can work without feeling stressed by administrative tasks. *Required to answer.*

| | Strongly disagree | Somewhat disagree | Neither agree nor disagree | Somewhat agree | Strongly agree |
|---|---|---|---|---|---|
| Without Tandem | ☐ | ☐ | ☐ | ☐ | ☐ |
| With Tandem | ☐ | ☐ | ☐ | ☐ | ☐ |

6. I feel fully present with the patient during the consultation. *Required to answer.*

| | Strongly disagree | Somewhat disagree | Neither agree nor disagree | Somewhat agree | Strongly agree |
|---|---|---|---|---|---|
| Without Tandem | ☐ | ☐ | ☐ | ☐ | ☐ |
| With Tandem | ☐ | ☐ | ☐ | ☐ | ☐ |

7. Compared to your previous way of working – does Tandem represent an improvement for your profession? *Required to answer. Single choice.*

- Yes
- No

8. Tandem is simple to use. *Required to answer. Single choice.*

- Yes
- No

9. I want to continue using Tandem. *Required to answer. Single choice.*

- Yes
- No

10. Do you recommend colleagues in your unit or at other units to use Tandem? *Required to answer. Single choice.*

- Yes
- No

---

### S2 Supplementary figures & tables

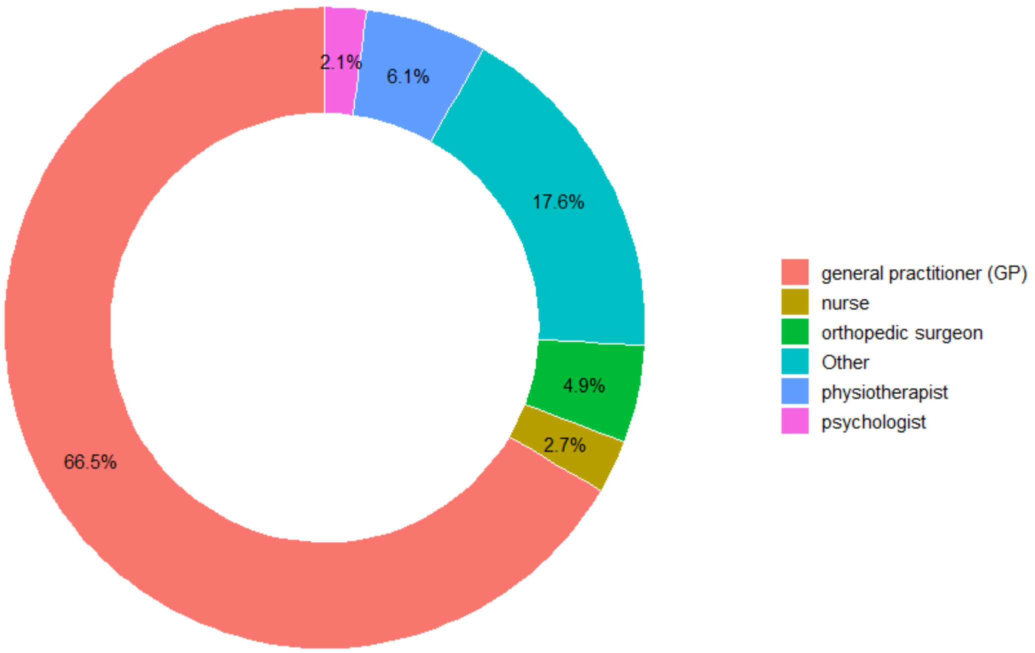


**Figure S1.** *Proportion of professions included in the study. The professions in "other" consist of less than 2% of the total professionals per profession.*

### ‘Distribution of edit times among edited notes’

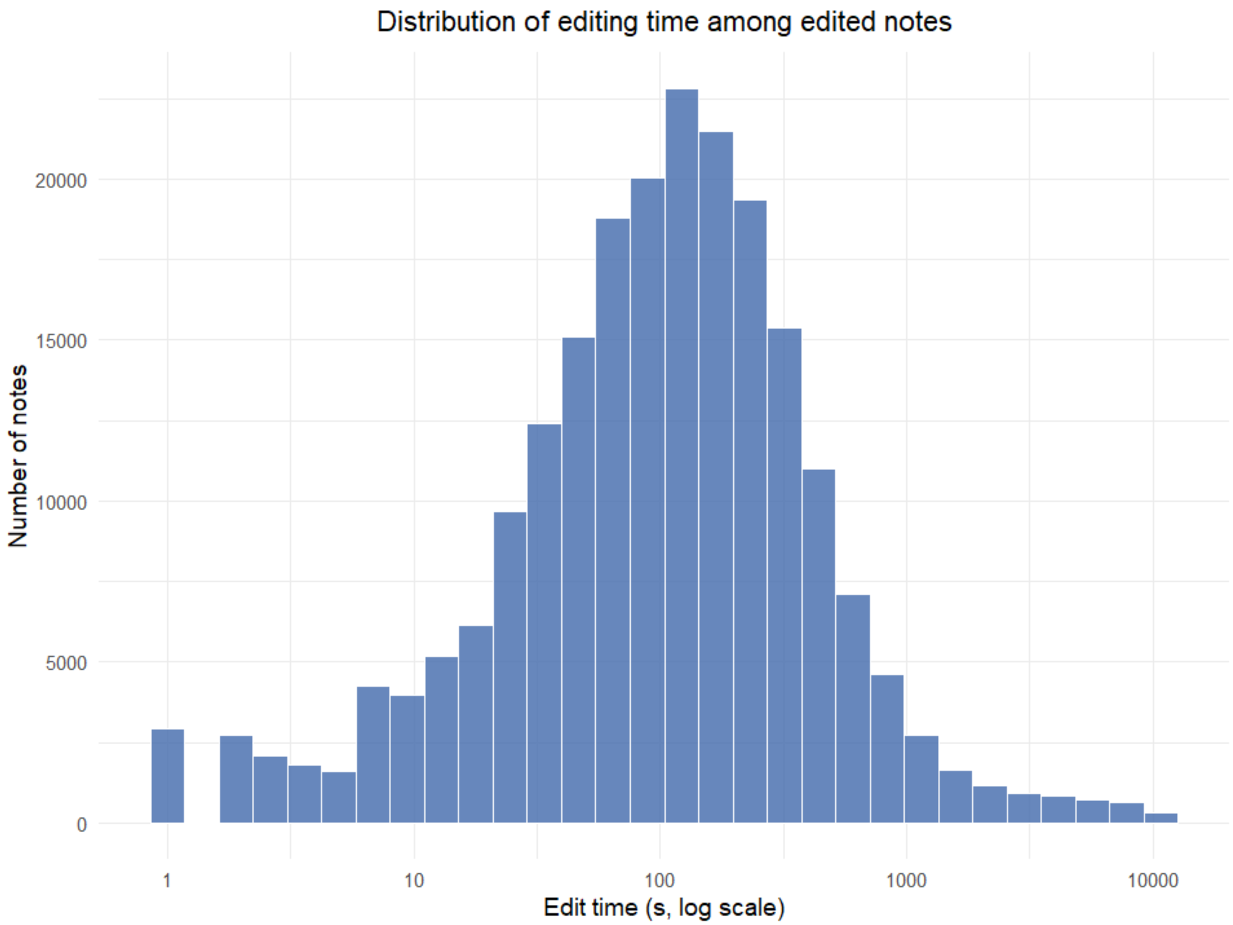


**Figure S2.** *Histogram with log-scaled axis.*
